# The science of child and adolescent mental health in Greece: a nationwide systematic review of the literature

**DOI:** 10.1101/2022.10.14.22281090

**Authors:** Anastasia Koumoula, Lauro Estivalete Marchionatti, Arthur Caye, Vasiliki Eirini Karagiorga, Panagiota Balikou, Katerina Lontou, Vicky Arkoulaki, André Simioni, Aspasia Serdari, Konstantinos Kotsis, Maria Basta, Efi Kapsimali, Andromachi Mitropoulou, Nikanthi Klavdianou, Domna Zeleni, Sotiria Mitroulaki, Anna Botzaki, Giorgos Gerostergios, Giorgos Samiotakis, Giorgos Moschos, Ioanna Giannopoulou, Katerina Papanikolaou, Katerina Aggeli, Nikolaos Scarmeas, Panagiotis Koulouvaris, Jill Emanuele, Kenneth Schuster, Eirini Karyotaki, Lily Kalikow, Katerina Pronoiti, Natan Pereira Gosmann, Julia Luiza Schafer, Kathleen R. Merikangas, Peter Szatmari, Pim Cuijpers, Katholiki Georgiades, Michael P. Milham, Mimi Corcoran, Sarah Burke, Harold Koplewicz, Giovanni Abrahão Salum

**Author notes:** **Corresponding author:** Giovanni Abrahão Salum^1,2,3^ MD PhD, Email address, Mailing address: Child Mind Institute. 101 E 56th St, New York, NY 10022, USA.

## Abstract

**Background:** Evidence-based information is essential to delivering effective mental health care, yet the extent and accessibility of the scientific literature are critical barriers for professionals and policymakers. To map the necessities and make validated resources accessible, we undertook a comprehensive analysis of scientific evidence on child and adolescent mental health in Greece.

**Methods:** This systematic review encompasses three research topics related to the mental health of children and adolescents in Greece: prevalence estimates, assessment instruments, and interventions. We searched Pubmed, Web of Science, PsycINFO, Google Scholar, and IATPOTEK from inception to December 16th, 2021. We included studies assessing the prevalence of conditions, reporting data on assessment tools, and experimental interventions. For each area, manuals informed data extraction and the methodological quality was ascertained using validated tools. This review was registered in protocols.io [68583].

**Outcomes:** We included 104 studies reporting 533 prevalence estimates, 223 studies informing data on 261 assessment instruments, and 34 intervention studies. We report the prevalence of conditions according to regions within the country. A repository of locally validated instruments and their psychometrics were compiled. An overview of interventions provided data on their effectiveness. The outcomes are made available in an interactive resource on-line [https://camhi.gr/en/systematic-review-tables/].

**Interpretation:** Scientific evidence on child and adolescent mental health in Greece has been cataloged and appraised. This timely and accessible compendium of up-to-date evidence offers valuable resources for clinical practice and policy making in Greece and may encourage similar assessments in other countries.

**Funding:** The Stavros Niarchos Foundation.

## Introduction

Evidence-based information is pivotal to the planning and delivery of effective mental healthcare.^1^ Prevalence surveys can identify frequent mental health conditions, assessing priorities of care and revealing at-risk groups. A pool of locally validated measurement instruments is key to mental health research and practice, as it allows professionals to screen, assess and monitor conditions in accordance with the best practices of measurement-based care.^2^ Empirical evidence on interventions informs strategies for effective allocations of resources. However, scientific literature is scattered and costly to assess, creating a ubiquitous barrier to evidence-based health delivery.^3–6^ Thus, moving science into action requires that evidence is available in a timely manner and has been synthesized and appraised according to well-established evidence-based principles.

Mental health services in Greece underwent a substantial modernization throughout the last few decades, including the implementation of child and adolescent specialized services and the development of a sectorized system.^7,8^ Nevertheless, a series of challenges remain for child and adolescent mental health care, mainly stemming from a lack of funding that was aggravated by the economic crisis.^9^ For instance, although policies pertaining to the sectorization of child and adolescent services were established,^10^ less than half of the planned services were created or implemented.^11^ There is still a shortage of child and adolescent psychiatrists in the public sector,^12,13^ particularly in rural areas as the majority of staff are located in the urban centers.^14^ Moreover, there are substantial gaps in epidemiological knowledge on mental health issues for the Greek general population,^15^ and mental health care provision has been understudied.^16^

The “Child and Adolescent Mental Health Initiative” (CAMHI) project is a five-year program that aims to enhance child and adolescent mental health care capacity and to help strengthen the infrastructure for the prevention, assessment, and treatment of mental health difficulties faced by children and adolescents across Greece. With the goal of facilitating the systematic uptake of evidence to inform policy and practice, we began this program by examining the scientific literature to map available resources and trace research priorities on child and adolescent mental health within the country. Herein, we report a comprehensive systematic review of the national scientific literature pertaining to child and adolescent mental health care in Greece. The review compiled, synthesized, and assessed prevalence surveys, validated assessment instruments, and interventions for mental health conditions among children and adolescents up to 18 years old. This accessible new compendium of up-to-date evidence-based information can offer valuable resources for both clinical practice and policy making in Greece and may encourage the undertaking of similar assessments in other countries.

## Methods

We followed the guidelines described by the Preferred Reporting Items for Systematic Reviews and Meta-Analysis (PRISMA) statement.^17^ The PRISMA checklist can be consulted in *Supplementary Table 1*. This study was registered in Protocols.io (number 68583),^18^ a platform that allows for registration after initiation of data extraction. For having an exceptionally broad scope as a nation-based systematic review, we anticipated that data extraction tables would require *ad hoc* adaptations and could only be traced after an overview of the magnitude and characteristics of findings. Therefore, we registered the protocol during the data extraction phase, when we defined appropriate methods for summarizing results.

### Search Strategy

Aiming at a comprehensive assessment of the Greek literature, a multi-step search strategy was employed targeting several electronic databases from inception to December 16th, 2021, without restrictions of language. First, we searched Pubmed, Web of Science and PsycINFO using English free terms that associate mental health conditions, children, and adolescents, and Greece (see *Supplementary Table 2* for the full queries). Then, we searched IATPOTEK database using corresponding Greek terms, aiming to reach local peer-reviewed literature. Duplicates were automatically removed using the software EPPI-Reviewer 4.0.^19^ As a complementary step, we searched Google Scholar using English and Greek terms. This is a crawler-based search engine that retrieves results that are exceedingly numerous to manage, and is recommended as an additional source of information for systematic reviews.^20,21^ Therefore, we used it to scan for studies that were not included in our primary set and as a source for gray literature. Two authors independently inspected results until reaching a mark of one hundred sequential studies that did not contain any novel inclusion. We also examined the reference list of studies for snow-balling inclusions and consulted local experts for additional references. All studies were uploaded to and managed with Rayyan,^22^ a platform designed to organize systematic reviews.

### Areas of systematic review

This review covers three research areas: prevalence estimates, assessment instruments, and interventions. Initially, we conducted an umbrella search to retrieve a set of studies that were screened and classified into those three research areas. Then, we carried out a three-arm review process, with distinct extraction and synthesis strategies for each topic.

### Inclusion criteria

#### Prevalence studies

We included studies reporting surveys on community-based, school-based, or other representative samples assessing the prevalence (lifetime, 12 months, and point prevalence) of mental health conditions (ascertained by clinical or structured interviews using ICD or DSM coding, or indicated by validated cut-offs on screening instruments) or the levels of mental health symptoms or of mental well-being/quality of life (using standardized instruments) among children and adolescents up to 18 years old in Greece.

Multi-country studies were included if data for Greek participants were separately presented. If the same dataset was reported in more than one study, we only included the most comprehensive or most recent manuscript. We included literature reviews for reference consulting purposes and dissertation thesis, academic letters, and book chapters were eligible.

We excluded surveys on clinical settings or non-representative samples (e.g., quality of life among leukemia patients). We also excluded conference abstracts.

#### Instrument studies

We included studies that developed, translated, validated, or solely applied instruments of screening, clinical assessment, or diagnosis of child and adolescent (up to 18 years old) mental health outcomes in Greece, regardless of the nature of the sample. Instruments developed for the adult or general population were included if the study applied it to our targeted population.

Multi-country studies were included if data for Greek participants were available. If an instrument was reported in more than one study, they were both included whenever presenting different information (e.g., distinct properties of psychometric validation). If two studies reported the same property (e.g., two internal consistency analyses), only the most powered study (i.e., highest sample size) was included. Literature reviews were included for reference consulting purposes. Dissertation thesis, academic letters, and book chapters were eligible if inclusion criteria were met. We excluded conference papers.

#### Intervention studies

We included studies reporting interventions for mental health conditions or for mental health promotion, including mental well-being/quality of life, targeting children and adolescents (up to 18 years old) in Greece. Any interventional design was eligible, from pre-post uncontrolled studies to randomized clinical trials. We also planned to include studies that aimed to translate or adapt interventions that proved effective in other settings, even without testing their effectiveness.

Multi-country studies were included if data for Greek participants were available. If two studies reported on the same trial, only the most recent one was included. Literature reviews were included for reference consulting purposes. Dissertation thesis, academic letters, and book chapters were eligible. We excluded papers on interventions that also included the adult population without discriminating data between age groups. In addition, we excluded conference abstracts.

### Screening process

During primary screening, two authors (AC, LEM) independently assessed results from databases searched with English terms. For databases searched with Greek terms, only one reviewer assessed the studies (VK). Studies were sorted into three areas of interest, and a specific study could be included in more than one area.

In the secondary screening, a single reviewer assessed full-text articles for final inclusion and extraction in each area. Any question of inclusion of a specific study was reviewed within the research team. Cross-group inclusions were allowed in this phase (for instance, if a study previously classified in the instrument area presented relevant information on prevalence).

### Data Extraction and Synthesis

#### Prevalence studies

We followed and adapted the extraction procedures of a systematic review and meta-analysis on the prevalence of child and adolescent mental health conditions.^23^ The following data were extracted: first author, year of publication, study description, region of which the study attempts to be representative, year of data collection, description of sampling/representativeness, age range, sex or gender distribution, details of screening and diagnostic sample procedures (screening sample size, screening response rate, screening instrument, screening informant, method for screening selection, diagnostic sample size, diagnostic response rate), diagnostic domain, condition or construct, assessment instrument, if instrument includes interview, informants, diagnostic criteria, if diagnosis requires functional impairment, definition of functional impairment, prevalence estimate and its standard deviation (SD) or 95% confidence interval (CI), and mean score and its standard deviation (SD). We evaluated risk of bias with a validated quality assessment tool for prevalence studies covering external validity, internal validity, analysis bias, and representativeness of the sample.^24^ Next, a summary table was derived to concisely present information. Finally, we created a synthesis table for aggregating information for each condition, including its lowest and highest prevalence estimate. In this table, only conditions with prevalence rates were included, as quality of life and other constructs counted only on mean scores obtained via assessment instruments. Due to the clinical heterogeneity across the studies, we did not perform a meta analysis of prevalence estimates.

#### Instrument studies

We followed the Consensus-based Standards for the Selection of Health Measurement Instruments (COSMIN) guidelines for creating data extraction sheets and for evaluating the strengths of findings and their methodological quality.^25^ All psychometric terms employed hereby are in accordance with the definition provided in the manual. Whenever an aspect of data extraction was not contemplated in the consensus instructions, we adapted a strategy for extraction after consulting scientific literature on the area.

The first section of the extraction protocol covered general information of the studies and instruments (first author, year of publication, name of the instrument, diagnostic domain, construct evaluated, original language, target population, informer/rater, recall period, number of items, response options, estimated time of application), sample description (sample size, age mean/range, percentage of females, diagnosed conditions, setting, response rate), and interpretability (proposed cut-off, percentage of missing items, floor and ceiling effects).

In the following section, we extracted information on study sampling and analytical procedures. First, a multi-choice field described study objectives (development, translation, validation, translation and validation, or application of an instrument). For studies on instrument translation, we included a checkbox to determine whether a back-and-forth translation procedure was undertaken. For instrument development studies, we ascertained the risk of bias of the items “development quality” and “content validity quality” in accordance with instructions of the manual. For studies that reported psychometric validation procedures (structural validity, internal consistency, cross-cultural validity, inter-rater reliability, test-retest reliability, measurement error, criterion validity, construct validity, or responsiveness), we also followed the manual and extracted three fields for each property: sample size for that procedure, methodological quality (very good, adequate, doubtful, inadequate, or not applicable), and results (see *Supplementary Table 3* for detailed extraction procedures).

Next, we derived a summary table to concisely present information on the psychometric properties of each instrument and study. Therein, we compiled data on instrument translation or development and a set of psychometric properties according to the manual coding: “+”, for sufficient; “-”, for insufficient; and “?”, for indeterminate. This rating followed COSMIN’s criteria, except for translation/development (see *Supplementary Table 4*) and responsiveness (see *Supplementary Table 5*). Finally, we created a synthesis table for aggregating the information on each instrument, rating psychometric properties as sufficient (“+”) whenever it was ascertained as such in at least one study.

#### Intervention studies

For building an extraction table, we followed and adapted the suggested extraction table of the Cochrane manual for systematic reviews of intervention studies.^26^ For each intervention study, the following data were extracted: first author, year of publication, sample size, experimental intervention, control intervention, diagnostic domain, target construct/disorder, primary outcome, primary outcome measurement, secondary outcome, secondary outcome measurement, sample description, age range/mean, percentage of females, ethnicity, inclusion criteria, exclusion criteria, recruitment method, allocation method, unit of allocation (individuals, clusters), duration of intervention, number of participants randomized by treatment arm, withdrawals and exclusions, other treatments received, subgroups, time points measured, person measuring/reporting, main results, funding, and conflicts of interest. Whenever possible, we calculated controlled and uncontrolled Cohen’s d effect sizes using reported means and standard deviations before and after the intervention using the command “escalc” from the package “meta” in the software R version 3.6.2.^27^ We evaluated methodological quality of studies through the revised version of the Cochrane risk-of-bias tool for randomized trials (RoB 2).^28^ For non-randomized designs, we used the Joanna Briggs Institute (JBI) checklist assessment tool.^29^ Next, a summary table was derived to concisely present information.

## Results

A total of 4,476 abstracts were retrieved in the umbrella search in Pubmed, Embase, PsyciNFO, and IATPOTEK databases, and another 1,074 were screened from Google Scholar. After de-duplication and full-text analysis, this yielded 102 publications in prevalence studies, 16 in instrument studies, and 27 in intervention studies. Reference list, expert consultation, and cross-group inclusions led to a further 2 inclusions in the prevalence domain, 207 in the instruments domain and seven in interventions. A final number of 104 prevalence studies, 223 instruments studies, and 34 intervention studies were included in the review. *Figure 1* details the screening procedures.

**Figure 1.**
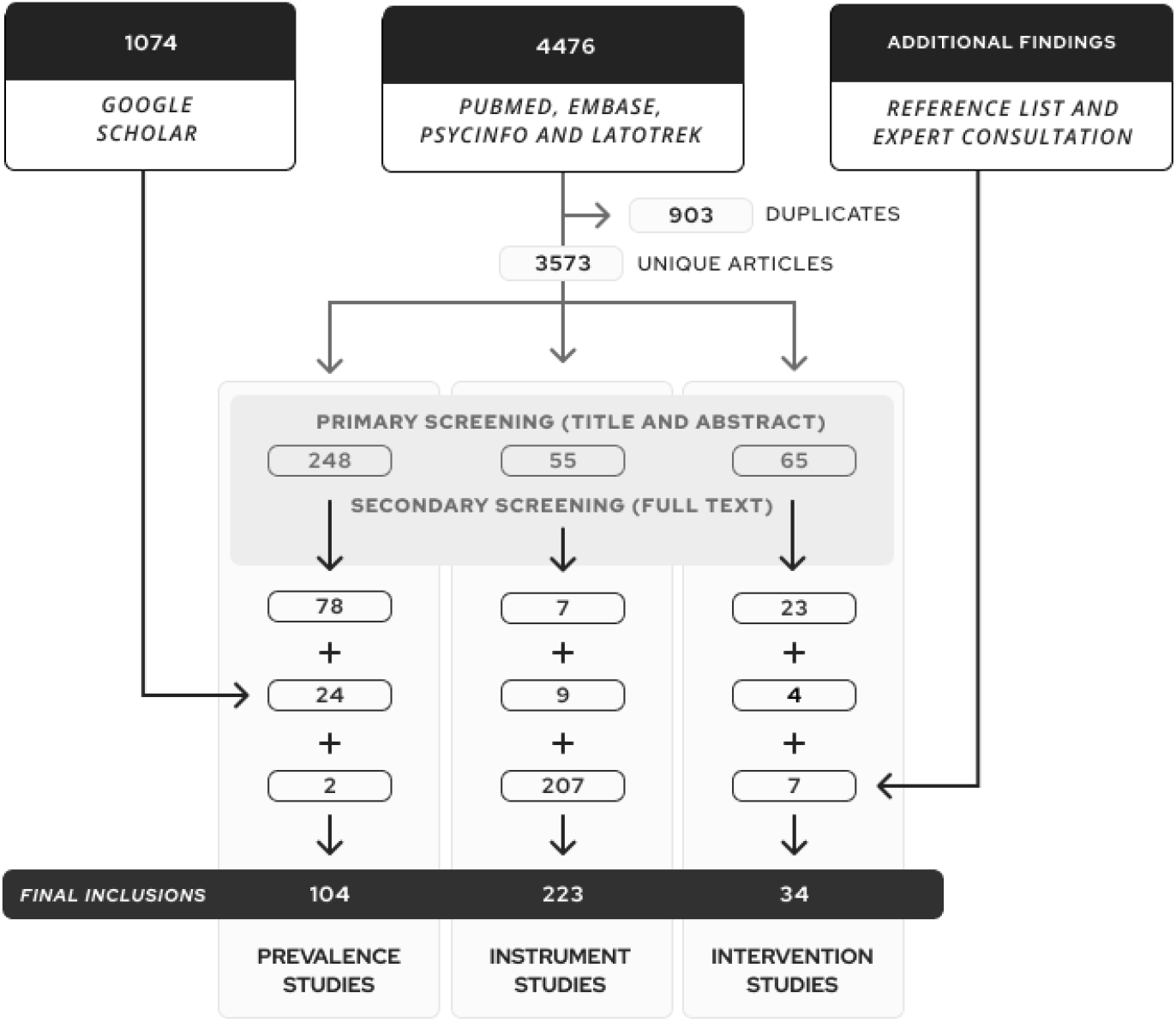
Flowchart describing the search and screening procedures

*Table 1* and *Table 2* summarize representative results for prevalence estimates and assessment tools, respectively. *Table 3* and *Table 4* describe all intervention studies. A free interactive dataset to navigate all findings across different levels of information is available online [https://camhi.gr/en/systematic-review-tables/]. The extraction table, summary table, synthesis table, and risk of bias assessment for each area are fully available in *Supplementary File 1*.

**Table 1.** Representative results for prevalence estimates on mental health conditions of children and adolescents in Greece

| Disorder or construct | Studies (N) | Age range | Years of data collection | Assessment instruments | Regions | Informants | Prevalence (lowest to highest) | References |
| --- | --- | --- | --- | --- | --- | --- | --- | --- |
| **Anxiety symptoms** | 1 (1.520) | 9 - 12 years | NA | SCAS-GR | Athens-Pireaus area | Self-report | 42·3% to 42·3% | Mellon (2007) |
| **Attention-deficit/hyperactivity disorder** | 4 (3.128) | 6 - 11 years | 2002, 2006-2007, 2009, 2011-2012 | CBCL, ADHD-IV, ADHD test, Lerner's Rating Scale of Student Behavior, Teacher's Report Form (TRF) | Athens, Attiki, Crete, Thessaloniki, Thessaly | Caregiver, Teacher | 2·6% to 6·5% | Skounti (2006)<br>Skounti (2010)<br>Papaioannou (2014)<br>Alemany (2021) |
| **Autism spectrum disorder** | 2 (183.187) | 6 - 11 years | 2011-2012, 2019 | Clinical diagnosis, CBCL | Crete, Nationwide | Caregiver, Clinician | 1·15% to 10·1% | Alemany (2021)<br>Thomaidis (2020) |
| **Bullying victimization** | 4 (3.679) | 12 - 18 years | 2003, 2008, 2016 | Revised Olweus Bully/Victim Questionnaire for Students-Senior, KIDSCREEN-52 | Crete, Thrace, Nationwide | Self-report | 12·4% to 30·5% | Koutra (2014)<br>Analitis (2009)<br>Serdari (2018)<br>Chudal (2021) |
| **Depressive disorder** | 3 (3.445) | 10 - 18 years | 1984, 2007-2008 | CES-D, CDI, CIS-R | Aetoloakarnania, Epirus, Island of Paros, Kallithea, Volos, Nationwide | Self-report | 5·7% to 30·5% | Madianos (1993)<br>Kleftaras (2006)<br>Magklara (2015) |
| **Internet addiction disorder** | 8 (9.152) | 12 - 18 years | 2007, 2008, 2009, 2010, 2011-2012 | Young Diagnostic Questionnaire, Internet Addiction Test (IAT), Young Internet Addiction Test (YIAT) | Athens, Attica, Chalkida, Island of Chios, Island of Kos, Korinthia, Peloponnese, Rhodes, Thessaly | Self-report | 0·8% to 16·1% | Siomos (2008)<br>Diakos (2010)<br>Fisoun (2012)<br>Trioda-Kyparissi (2011)<br>Tsitsika (2011)<br>Siomos (2012)<br>Stravopoulos (2013)<br>Stravopoulos (2018) |
| **Insomnia** | 2 (2.908) | 13 - 18 years | NA | Athens Insomnia Scale (AIS) | Greater Athens, Thessaly | Self-report | 11·44% to 56·5% | Lazaratou (2005)<br>Siomos (2010) |
| **Alcohol use (regular use)** | 5 (4.020) | 11 - 18 years | 2003-2004, 2005-2006, 2008 | NA | Crete, Karlovasi, Ioannina, Thessalonik, Nationwide | Self-report | 10·5% to 51·9% | Elisaf (1996)<br>Arvanitidou (2007)<br>Farhat (2012)<br>Koutra (2014)<br>Garoufi (2017) |
| **Cannabis (lifetime)** | 4 (24.637) | 12 - 17 years | 1984, 1993, 1998, 1999, 2001-2002, 2003, 2004, 2007, 2011, 2015 | NA | Nationwide | Self-report | 2·4% to 12·1% | Kokkevi (2006)<br>Kokkevi (2007)<br>Kokkevi (2007)b<br>Kokkevi (2016) |
| **Smoking tobacco (regular use)** | 15 (76.393) | 12 - 18 years | 1984, 1993, 1998, 1999, 2001 - 2002, 2003, 2003 - 2004, 2004 - 2006, 2005, 2007, 2008, 2011, 2015 | WHO-GYTS | Crete, Ioannina, Karlovasi, Kos, Thessaloniki, Nationwide | Self-report | 5·4% to 38·1% | Kokkevi (1991)<br>Elisaf (1996)<br>Kokkevi (2000)<br>Francis (2007)<br>Kyrlesi (2007)<br>Kokkevi (2007)<br>Arvanitidou (2008)<br>Heras (2008)<br>Lazuras (2010)<br>Giannakopoulos (2010)<br>Soteriades (2011)<br>Kokkevi (2012)<br>Koutra (2014)<br>Kokkevi (2016)<br>Garoufi (2017) |
| **Suicide attempt** | 3 (70.549) | 12 - 18 years | 1984, 1993, 1998, 2003, 2007, 2011 | CES-D | Nationwide | Self-report | 1·9% to 15·2% | Madianos (1993)<br>Kokkevi (2010)<br>Kokkevi (2014) |
| **Suicide mortality** | 4 (NA) | 0 - 18 years | 1980 - 1995, 1994-1996, 2000 - 2009 | Register | Nationwide | Register | 0% to 0·01% | Beratis (1991)<br>Zacharakis (1998)<br>Rutz (2004)<br>Bacopoulou (2015) |
| **Sexual violence** | 1 (10.451) | 11 - 16 years | NA | ICAST-C | Nationwide | Self-report | 15·86% to 15·86% | Nikolaidis (2018) |

**Note:** Results were selected from the full dataset considering the relevance of mental health conditions and the number of studies reporting estimates. *Supplementary File 1* presents the entire set of prevalence estimates. **Abbreviations:** not available (NA).

**Table 2.**
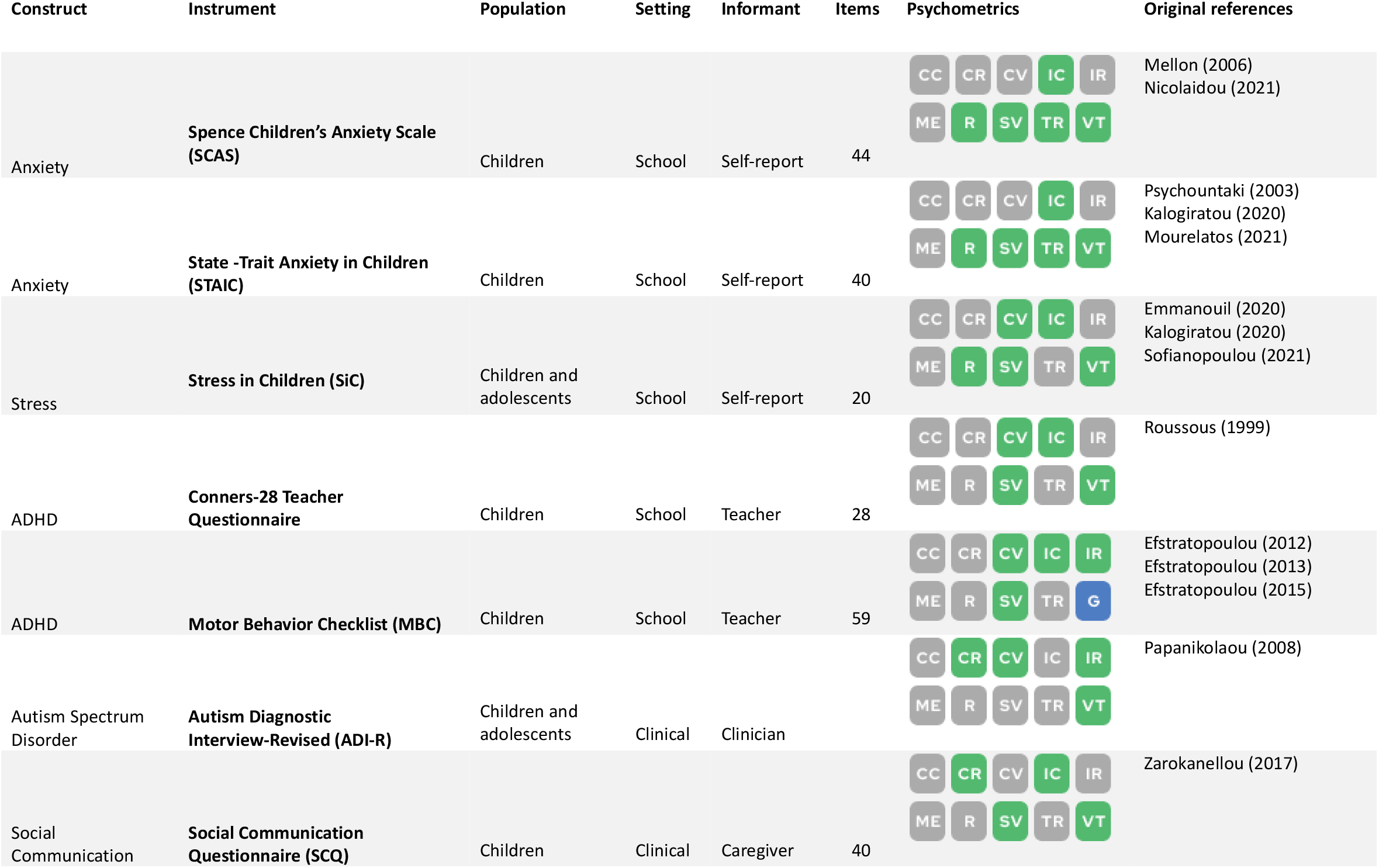
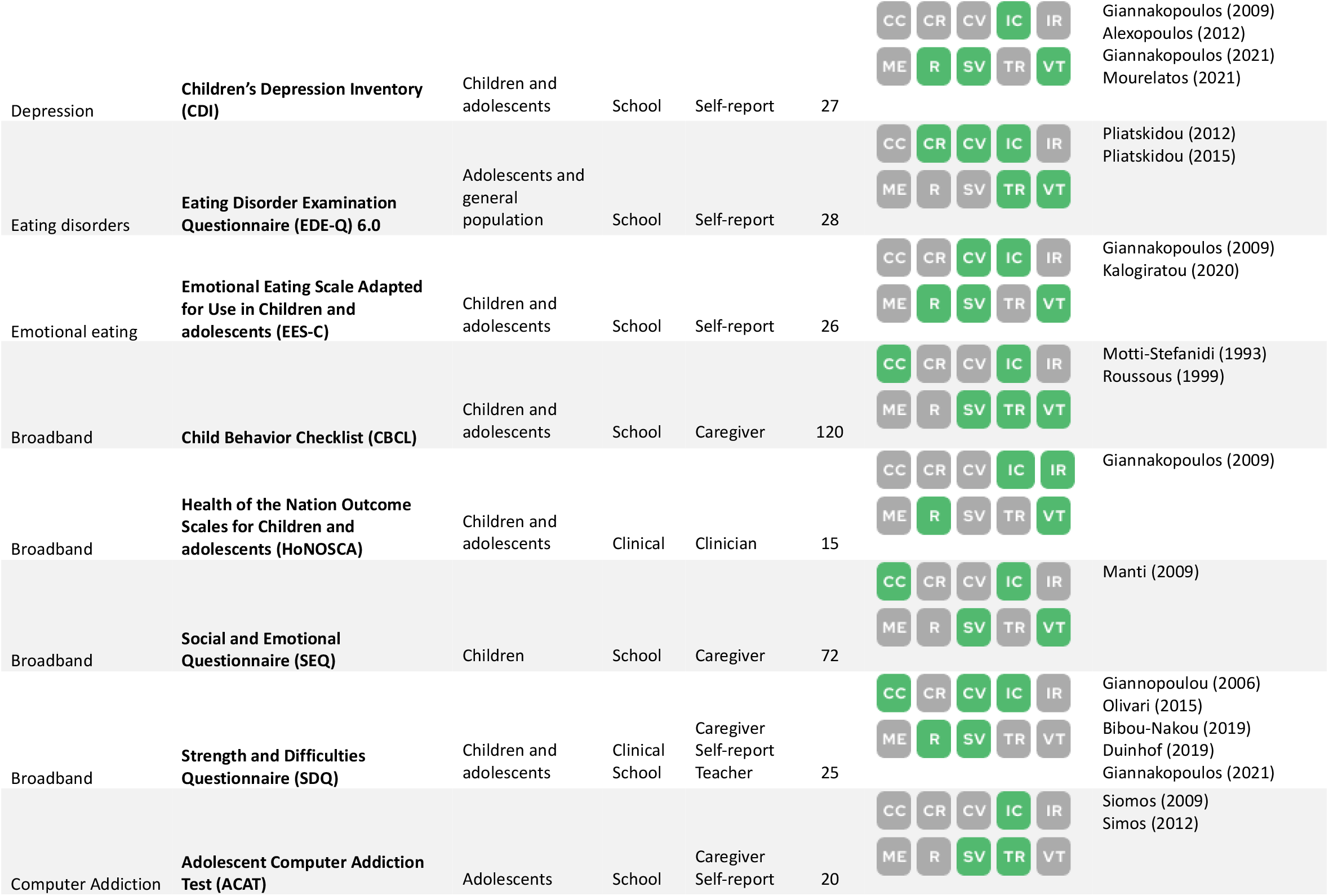
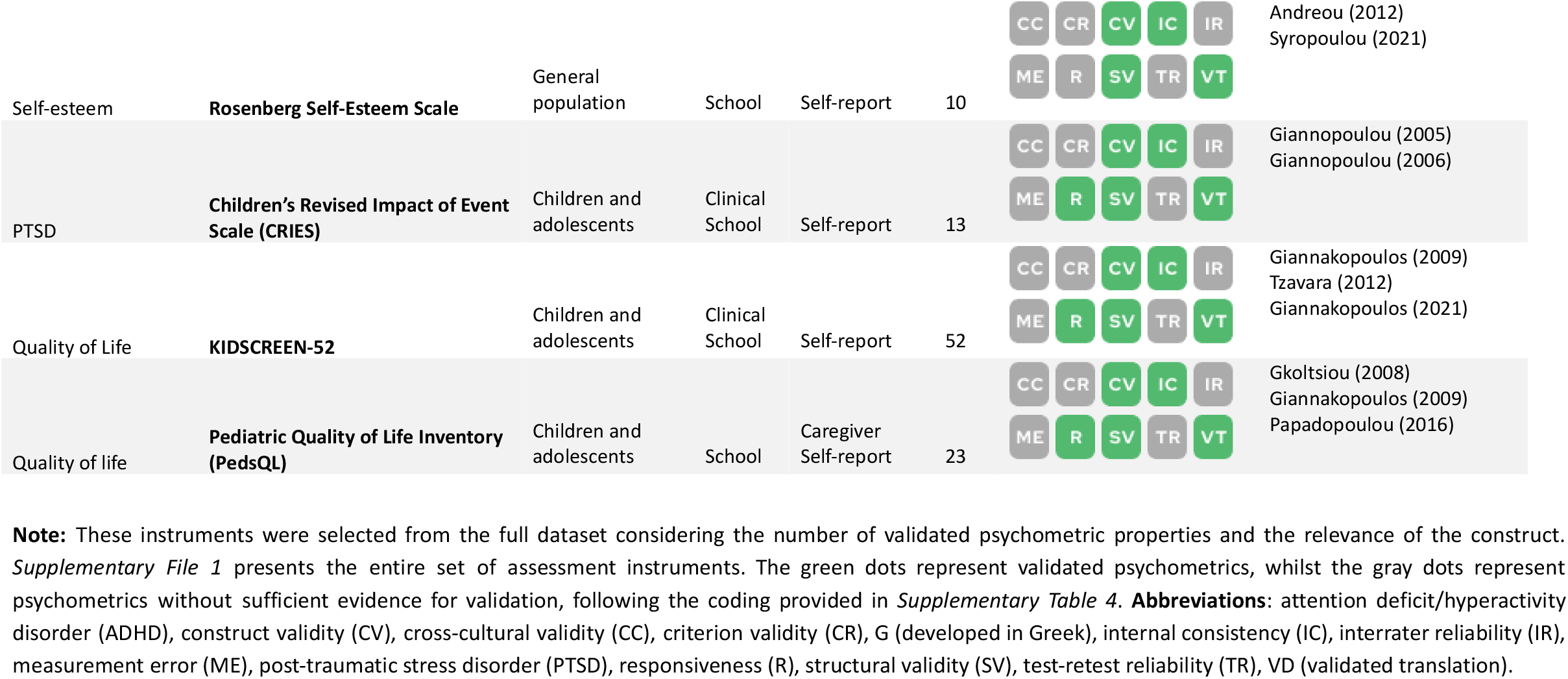
Selected results for assessment instruments on mental health outcomes validated for children or adolescents in Greece

**Table 3.** Experimental interventions on child and adolescent mental health conducted in Greece (controlled designs)

| First author (year) | Intervention | Primary outcome | Participants | Design | Demonstrated efficacy? | Effect size (controlled uncontrolled) |
| --- | --- | --- | --- | --- | --- | --- |
| Sofianopoulou (2021) | Diaphragmatic breathing and progressive muscular relaxation training | Stress scores | 52 | RCT | Yes | 1·29 (0·16)<br>1·47 (0·11) |
| Lappa (2021) | Music and dramatic art education | Social behavior | 30 | Quasi-experimental | Yes | 0·41<br>0·28 |
| Giannakopoulos (2021) | Family Talk Intervention group | Emotional/behavioral problems<br>Prosocial behavior<br>Anxiety/depression symptoms | 62 | RCT | No | 0·01 – 0·16<br>0·76 – 0·86 |
| Nicolaidou (2021) | Web-based learning environment on resilience | Anxiety scores | 41 | Quasi-experimental | No | -0·21 - 0·58<br>-0·12 - 0·42 |
| Foka (2020) | Strengths for the Journey (SFJ) group program | Overall well-being | 72 | Quasi-experimental | Yes | 1·79 (0·16)<br>2·18 (0·12) |
| Kalogiratos (2020) | Pythagorean Self Awareness Intervention (PSAI) | Emotional eating scores | 55 | RCT | Yes | 1·00 (0·15)<br>0·94 (0·08) |
| Sourvinos (2020) | Aquatic Speech and Language Therapy (ASLT) | Expressive and receptive vocabularies | 16 | Quasi-experimental | Yes | 1·45 (0·35 - 2·56) /<br>1·92 (0·74 – 3·10)<br>NA |
| Makrygianni (2017) | ABA Intervention Programs<br>Type-TEACCH Programs<br>Eclectic programs | Autism behavior | 30 | Quasi-experimental | No | NA<br>NA |
| Koutsoura (2017) | Psychoeducational group program based on forgiveness | Symptoms of depression | 42 | RCT | Yes | 1·15<br>2·46 |
| Hatzichristou (2017) | Teachers' training program for the promotion of resilience and positive school climate | Teachers' resilience | 606 | Quasi-experimental | Yes | 0·25 - 0·373<br>0·0 - 0·058 |
| Vassilopoulos (2015) | Attribution Modification Program | Aggressive behavior | 34 | RCT | Yes | 0·39 (0·18)<br>0·21 (0·10) |
| Vassilopoulos (2014) | Interpretation of cognitive bias training | Negative interpretation | 94 | RCT | Yes | NA<br>NA |
| Chiou (2013) | Mental health promotion intervention program named "I Can Problem Solve" | Cognitive skills<br>Social skills<br>Problem Behaviors | 151 | Quasi-experimental | Yes | 0·55 - 1·34<br>NA |
| Economou (2012) | Group-based educational talk | Beliefs and attitudes towards Schizophrenia | 1081 | RCT | Yes | NA<br>1·45 (0·005) |
| Makrygianni (2011) | ABA Intervention Programs | Progress on developmental domains | 21 | Quasi-experimental | Yes | NA<br>0·14 - 0·8 |
| Economou (2011) | A semi-structured anti-stigma program with group dynamics | Beliefs and attitudes towards Schizophrenia | 616 | Quasi-experimental | Yes | NA<br>0·17 – 1·72 |
| Vassilopoulos (2009) | Interpretation of cognitive bias training | Social anxiety scores | 43 | RCT | Yes | 1·87 (0·21)<br>1·73 (0·14) |
| Hatzichristou (2009) | Positive psychology intervention program | Problem Solving<br>Social and Emotional skills<br>Resilience | 1319 | Quasi-experimental | Yes | NA |
| Andreou (2007) | Bullying Awareness Program within schools | Bullying behavior | 454 | Quasi-experimental | No | 0·38 (0·001)<br>0·11 (0·01) |
| Zafiropoulou (2007) | Positive Emotions Program for Preschoolers | Ability to identify thoughts and emotions | 20 | RCT | Yes | 0·35 (0·33) / 0·29 (0·31)<br>0·74 (0·18) / 0·50 (0·16) |
| Tarnanas (2004) | Virtual Reality Cognitive-behavioral treatment | Hazard adjustments | 209 | Quasi-experimental | Yes | NA<br>1·12 |
| Koumi (2001) | Working groups for the production of audio-visual material with anti-smoking messages | Smoking behavior | 657 | Quasi-experimental | Yes | 0·39 (0·007)<br>NA |

**Abbreviations:** Applied behavior analysis (ABA), randomized controlled trial (RCT)

**Table 4.**
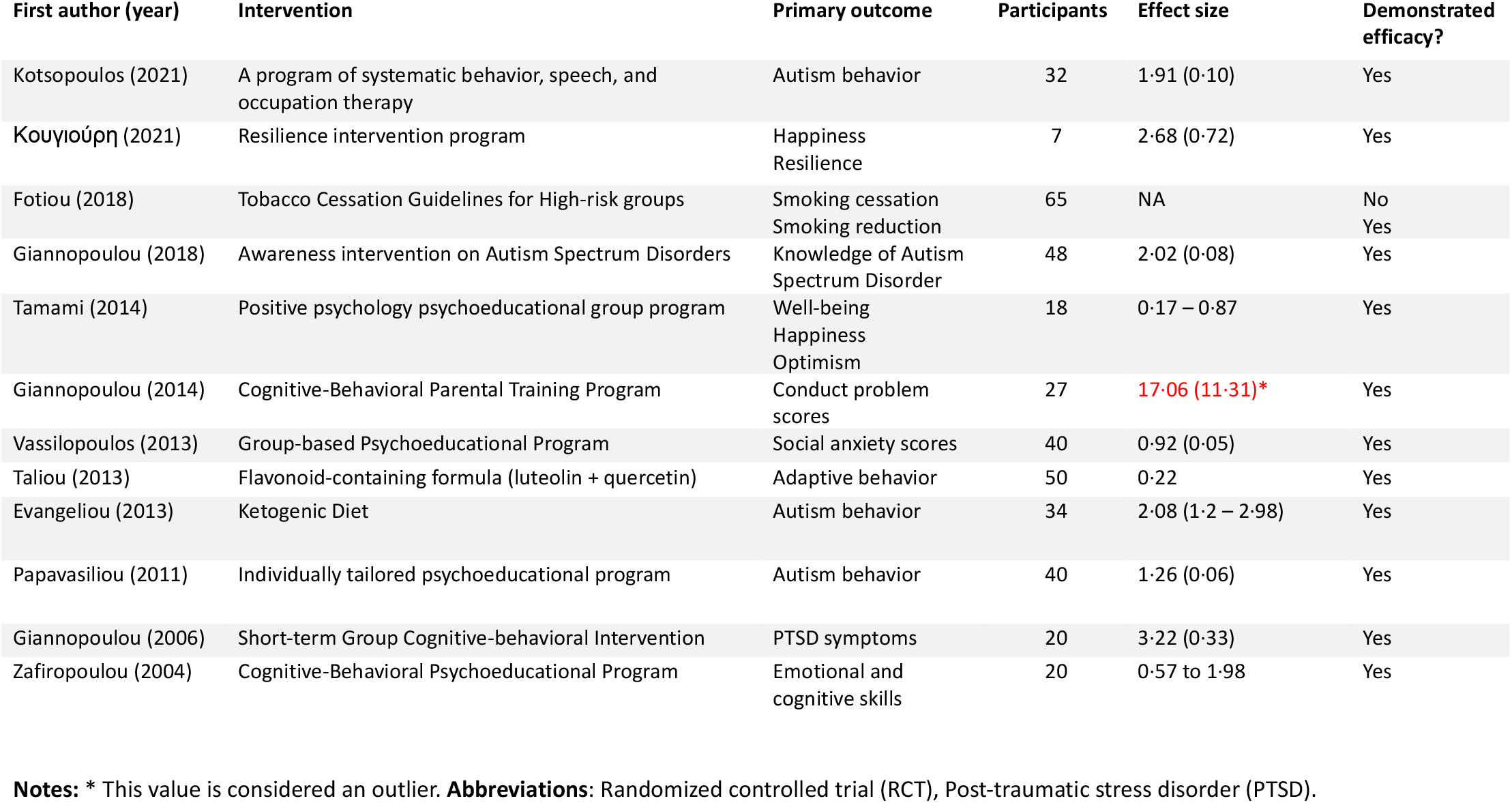
Experimental interventions on child and adolescent mental health conducted in Greece (uncontrolled designs)

*Figure 2* displays the number of studies in each research topic according to diagnostic domains, representing areas of concentration of scientific production. In this sense, general psychopathology was the domain counting on the most number of articles, as 17 prevalence studies reported data on constructs such as internalizing symptoms and emotional functioning, and another 34 instrument studies reported instruments that assess symptoms from multiple domains or general constructs. Next, a significant focus of scientific papers was neurodevelopment, with three studies on prevalence estimates, 47 on assessment tools, and one on experimental interventions. This is followed by anxiety disorders, quality of life, substance use disorders, and attention deficit/hyperactivity disorders, to name a few. Domains such as violence and neglect, sleep disorders, and obsessive compulsive disorders counted on very few reports across all research areas. No reports were found for domains such as bipolar disorder and psychotic disorders.

**Figure 2.**
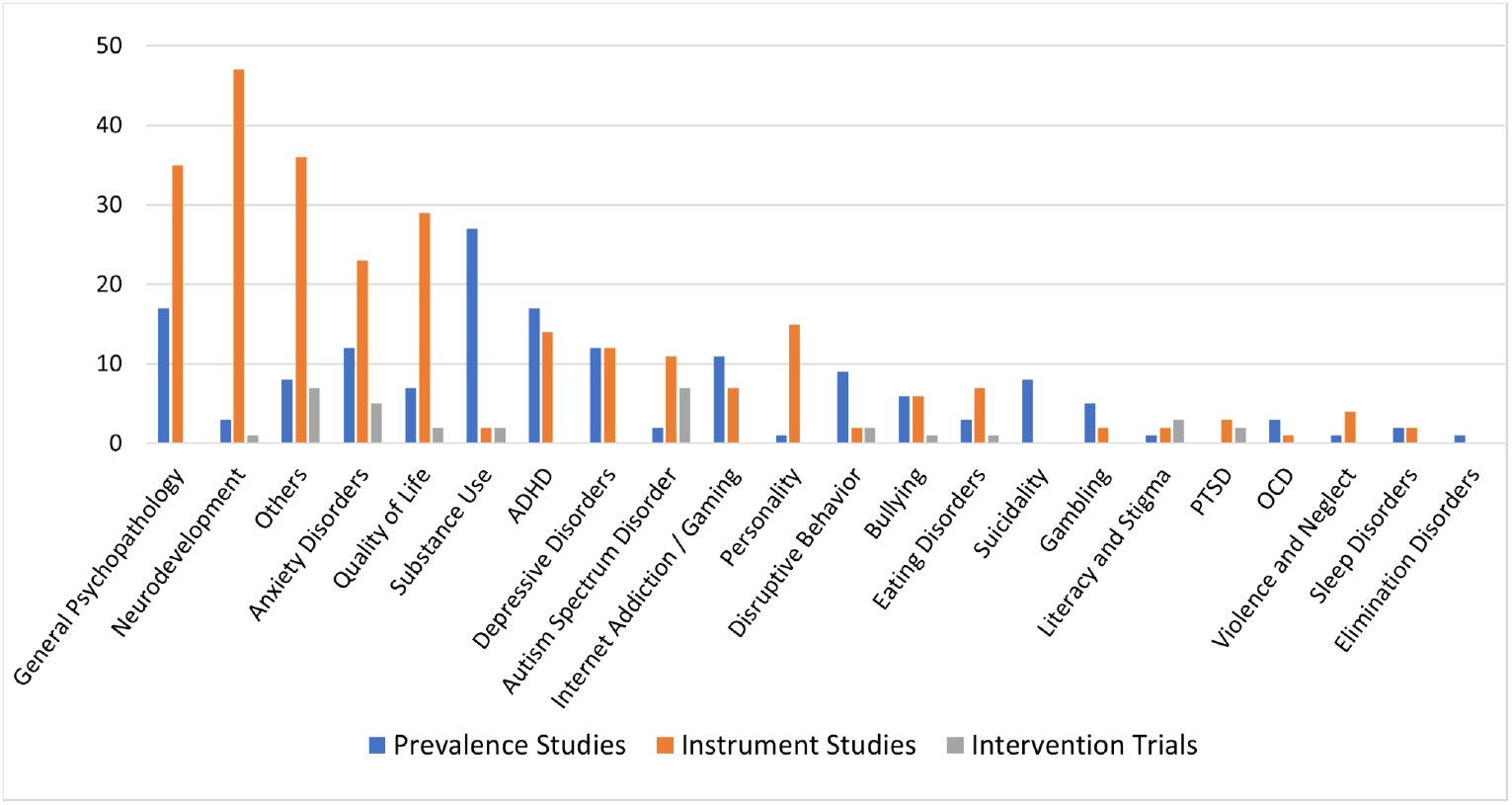
Areas of concentration of scientific production on child and adolescent mental health in Greece. **Abbreviations**: attention deficit/hyperactivity disorder (ADHD), obsessive-compulsive disorder (OCD), post-traumatic stress disorder (PTSD).

### Prevalence estimates

A total of 104 studies reported 533 estimates of the prevalence of mental health conditions, levels of mental health symptoms, or levels of quality of life/well-being. Most studies used scale cut-off points to ascertain the presence/absence of a mental health condition, which can greatly overestimate prevalence in comparison to gold-standard assessments such as clinical interviews. Only four estimates relied on clinical diagnosis or structured interviews. Most studies had their methodological quality rated as good, with some considered very good and a few presenting quality concerns. Tobacco use was the most commonly reported prevalence estimate, with 15 studies pointing to rates of regular smoking from 5·4% to 38·1% in samples aged 12 to 18 years of varying regions of the country (Crete, Ioannina, Karlovasi, Kos, Thessaloniki, Nationwide). The prevalence for internet addiction disorder was reported in eight studies, rates ranging from 0·8% to 16·1% in samples from 12 to 18 years old surveyed between 2007 and 2012. Another frequently reported condition was attention-deficit/hyperactivity disorder, and estimates from four studies point to prevalence rates from 2·6% to 6·5% in samples aged 6 to 11 years old surveyed using instruments rated by caregivers or teachers. For depressive disorder, three studies pointed to estimates from 5·7% to 30·5% in samples from 10 to 18 years old, with the last survey taking place in 2008. Autism spectrum disorder estimates were available in two studies, reporting prevalences from 1·15% to 10·1% in samples aged 6 to 11, which were obtained using caregiver-rated screening tools and also clinical diagnosis. There were no prevalence estimates for diagnosis such as generalized anxiety disorder or psychotic disorders, but one study reported a prevalence of 42·3% for anxiety symptoms in a 9 to 12 year-old sample from the Athens-Piraeus area. Studies were mostly concentrated in the regions of Attica, Crete, and Thessaloniki.

### Assessment instruments

A total of 223 studies reported information on 261 instruments that measure mental health outcomes. Specific domains with the highest number of available instruments were: neurodevelopment (52 instruments), general psychopathology (28 tools classified as broadband mental health instruments or measuring general constructs), anxiety disorders (17 instruments), and autism spectrum disorders (13 instruments). There was a scarcity of instruments for the domains of substance use, child abuse, and psychotic disorders. The most studied instrument was the Strength and Difficulties Questionnaire (SDQ), with information reported on five positively rated psychometric properties taken from five studies. The majority of instruments were translated from their original English version using a validated back-and-forth procedure, and 48 measurement tools were reported to be originally developed in Greek. Psychometric properties like internal consistency were considered sufficient and counted on adequate methodological quality for most studies, whilst others like cross-cultural validity, criterion validity, and measurement error were seldom reported or had significant risk of bias. Most studies had psychometric validation of some, but not most, properties.

### Intervention studies

A total of 34 studies reported different interventions conducted in clinical trials. Most frequently, they evaluated psychosocial group interventions (e.g. anti-stigma programs at a school, anti-smoking education, problem-solving skills, psychoeducational programs), but there were also a few individual or mixed interventions, one pharmacological therapy (a formulation with compounds from the *Sophora japonica* for adaptive behavior in autism), and one dietary intervention (ketogenic diet for Autism Spectrum Disorder). Twelve studies applied uncontrolled before-after designs, 13 studies used quasi-experimental designs, and nine studies were randomized clinical trials. Risk of bias for all studies was rated as high. There were no studies that aimed to validate interventions that proved effective in other settings.

## Discussion

This is a comprehensive systematic review of the scientific literature on the mental health of children and adolescents in Greece, which compiled and assessed studies reporting prevalence estimates, instrument assessment tools, and intervention trials. We compiled data on 533 prevalence estimates or level of symptoms for over 79 conditions or constructs in varying regions from Greece, mapped resources of 261 locally validated assessment tools, and provided an overall picture of 34 interventions reported in clinical trials in the country. This landscape analysis revealed prevalent conditions according to localities and uncovered conditions for which data are lacking, as well as regions that have not been sufficiently contemplated by research. It also provided a map of locally-validated instruments, alongside their characteristics and psychometric properties. Finally, the landscape analysis brought together a set of locally-studied interventions, revealing data on their effectiveness, methodological strengths, and weaknesses, and elucidated needed directions for upcoming research.

This work can be understood within the framework of implementation science, as it aims to address the well-documented gap between research and mental health practice.^30^ A pivotal step in addressing this challenge is to make scientific data easily available and appraised according to evidence-based principles, as real-world scenarios require up-to-date and accessible information for decision-making. This review provides such a compendium of data and information about available tools and estimates, which can readily inform professionals, policymakers, and other stakeholders in the fields of mental health assistance and research. Using the freely available interactive tool we have created to navigate this data, mental health practitioners and researchers can consult these resources to find locally-validated instruments that most suit their needs in both research and clinical settings, with options to filter it according to psychometrics, informants, or age group. Policy makers could also turn to this dataset for validated guidance when planning mental health programs, as the prevalence estimates of mental health conditions indicate targets for interventions, helping to establish priorities for care and to guide the allocation of resources. To the best of our knowledge, this is the first initiative to undertake a comprehensive national review and assessment of the literature on child and adolescent mental health.

This systematic review has many strengths. It has a broad scope with a comprehensive search strategy, including a range of databases, snowballing inclusions, and expert consultation, without restrictions of time or language. It also follows appropriate evidence synthesis guidelines for the three domains (i.e., prevalence estimates, assessment instruments, and interventions), rigorously inspecting studies according to manuals like the Cochrane, COSMIN, or established evidence-based practices. Furthermore, it synthesizes a significant amount of results in an accessible manner for consultation.

Throughout the process, we also encountered a number of limitations. As an overall issue, we could not a priori define the best data synthesis strategy, which could only be traced after visualization of data extraction, delaying protocol registration up to this point. In the instruments area, the review scope is particularly challenging since many translations and validations exist in sources that may not be captured by our search strategy (books, conferences, manuals, or developer websites). We tried to address this by extensively consulting the reference list from studies and taking recommendations from experts in the area, which accounted for a significant number of inclusions that outnumbered the inclusions from our primary dataset.

In conclusion, this landscape analysis can serve as a critical tool in bridging the gap between research and practice in Greece, as clinicians and policymakers can easily access tools and data to inform their practice and priorities, and which may serve to encourage similar nationwide studies beyond Greece.

## Data Availability

All data produced in the present work are contained in the manuscript

## Acknowledgments

The authors would like to thank the Stavros Niarchos Foundation (SNF) for funding the SNF-CMI Child and Adolescent Mental Health Initiative and SNF’s Co-President Andreas C. Dracopoulos for his leadership in creating, launching, and supporting the project. We would also like to thank Ms. Elianna Konialis, Ms. Dimitra Moustaka and Mr. Panos Papoulias for their critical role in multiple steps of the conceptualization and implementation of the SNF-CMI Child and Adolescent Mental Health Initiative. We also thank Samanta Duarte for designing the graphical representations included in this paper.

